# Is it time to use machine learning survival algorithms for survival and risk factors prediction instead of Cox proportional hazard regression? A comparative population-based study

**DOI:** 10.1101/2021.11.20.21266627

**Authors:** Sara Morsy, Truong Hong Hieu, Abdelrahman M Makram, Osama Gamal Hassan, Nguyen Tran Minh Duc, Ahmad Helmy Zayan, Le-Dong Nhat-Nam, Nguyen Tien Huy

## Abstract

**Purpose:** Applying machine learning in medical statistics offers more accurate prediction models. In this paper, we aimed to compare the performance of the Cox Proportional Hazard model (CPH), Classification and Regression Trees (CART), and Random Survival Forest (RSF) in short-, and long-term prediction in glioblastoma patients.

**Methods:** We extracted glioblastoma cancer data from the Surveillance, Epidemiology, and End Results database (SEER). We used the CPH, CART, and RSF for the prediction of 1- to 10-year survival probabilities. The Brier Score for each duration was calculated, and the model with the least score was considered the most accurate.

**Results:** The cohort included 26473 glioblastoma patients divided into two groups: training (n = 18538) and validation set (n = 7935). The average survival duration was seven months. For the short- and long-term predictions, RSF was the best algorithm followed by CPH and CART.

**Conclusion:** For big data, RSF was found to have the highest accuracy and best performance. Using the accurate statistical model for survival prediction and prognostic factors determination will help the care of cancer patients. However, more developments of the R packages are needed to allow more illustrations of the effect of each covariate on the survival probability.

## Introduction

Malignant brain tumors are among the most formidable types of cancer, with their poor prognosis and the direct influence on cognitive functions, working ability, quality of life [1]. In 2010, according to the prevalence estimate, nearly 200000 patients were diagnosed with primary malignant brain tumors in the United States [2]. Among all the primary malignant brain tumors, malignant gliomas are the most common type with 80% of patients and an annual incidence of 5.26 per 100 000 population, which also means 17000 new cases diagnosed per year [3]. This disease, however rare in children, may present at any age, but it peaks in the sixth through eighth decades of life. Moreover, the number of patients is expected to increase with the aging of the population [3-5].

Glioblastoma (GB) is the most frequent subtype that comprises 51% of all gliomas [6]. Because of its location in the brain, aggressiveness, and low survival duration, GB is well-known as one of the most lethal cancers [7]. The common symptoms in the last month of life include seizures, headache, drowsiness, dysphagia, and eventually death rattle, agitation, and delirium. In the last stage of the disease, patients need appropriate palliative care to allow them to experience a peaceful death despite their severe symptoms [7]. According to 2016 CNS WHO classification, glioblastomas are separated into glioblastoma, IDH-wildtype (presenting in 90 % of cases), which conforms most frequently with the clinically defined primary and prevails in patients over 55 years of age [1, 8]; glioblastoma, IDH-mutant (in about 10 % of cases), which conforms closely to secondary glioblastoma and tendentiously appears in younger patients [8]; and another is glioblastoma, NOS (not otherwise specified), a diagnosis for those tumors which full IDH evaluation cannot be performed [1].

In research, especially epidemiological topics, scientists often encounter multilevel or hierarchical data, such as evaluating the potential characteristics of patients, hospitals, and regions related to the risk of death in those patients with glioblastoma during a specified duration. Survival analysis, thus, refers to methods for the analysis of data in which the outcome demonstrates the time to the onset or occurrence of a targeted event. This method of analysis has the characteristic of censorship: the event may not occur for all subjects before the completion of the study and, at the end of the study, those event-free subjects are said to be censored [9, 10].

The most basic method for survival analysis is survival tables [9, 10]. The time is divided into intervals. For each interval, the count and proportion of each living, censored and death cases are calculated. The most widely used method is the Cox Proportional Hazard regression model or approach (CPH). It estimates the magnitude of the risk of death and its confidence interval. It is used for multiple analyses of survival time data. It is considered a multiple linear regression analysis. CPH analysis depends on the assumption of the proportionality of survival time data [10]. The results are hazard ratios that estimate the probability of an event at a specific type. One of the most common cons of CPH is the convergence or the divergence of the model. This occurs if the assumption of proportionality is not fulfilled or in the presence of many covariables that are not important. It is also reported that CPH analysis had overfitting problems, which means that it describes the random error instead of examining the relationship between the variables [10].

With the development of different machine learning algorithms, new algorithms were used to deal with different limitations and problems in biomedical research. In survival analysis, many algorithms have been used. One of them is recursive partitioning algorithms. These algorithms include Random Survival Forests (RSF) and Classification and Regression Trees (CART) [11-13].

The CART algorithms have gained popularity over time due to easy implementation and interpretability. It is based upon using important variables to split the data into many nodes representing the predictors. Impurity parameters are used to identify whether to select the splitting variable and whether to continue or stop the splitting. In each node, there is a strong association between splitting variables and the response variables evidenced by the highest impurity reduction [14, 15].

Meanwhile, an RSF is an algorithm of an ensemble of trees; it is the average prediction of all trees that would produce a more accurate prediction. It is a robust algorithm against overfitting and resistant to outliers and high dimensionality data. It is a nonparametric method that can be used on any variables regardless of the distribution they follow [12, 13, 15].

In this study, we aim to compare the performance of the Cox Proportional Hazard approach (CPH), CART, and Random Survival Forest (RSF) in short-, and long-term prediction in glioblastoma patients.

## Methods

### Data collection

We extracted the data of cases diagnosed with glioblastoma as reported in the histology recode of brain grouping in the SEER database. We used the last version of the published US research data (1975 – 2018) released on April 2021 [16].

This included glioblastoma, NOS (9440/3), Gliosarcoma (9442/3), and giant cell glioblastoma (9441/3). SEER*stat 8.3.9.2 was used to extract the data [17]. We only included cases that died due to the tumor itself and survival time more than zero. Cases with a survival time of zero were excluded.

Based on literature, long-term survival was defined in literature as survival of glioblastoma patients more than two years [18].

### Statistical analysis

The categorical variables were expressed in percentage while the continuous variables are expressed using mean and standard deviation if the data are normally distributed; otherwise, median, and interquartile range. The dataset was divided randomly into training groups with 70% of the cases and a test set with 30% of the dataset using the caret package. In the training set, missing data were imputed using the K nearest neighbors (KNN) algorithm with the number of neighbors equal to 3. The imputation was conducted in R using the VIM package [19]. For the validation set, we did sensitivity analysis where we compared the performance results between

i Imputation of the missing data of the validation set using KNN
ii Omitting the missing data from the validation set

Three survival algorithms were performed and compared based on an Brier Score (BS) for survival analysis on intervals of 12 months to compare the performance of the models for short- and long-term prediction. Univariable Cox Proportional Hazard regression analysis (CPH) was first developed to detect the significant covariables which were used in the multivariable analysis. Then, the accuracy of the model was assessed on the validation set using Brier score.

Random Survival Forests (RSF) were applied using the randomforestSRC package in R [20, 21]. We used 500 trees with three variables to split at each node. The variable importance was detected using the permutation method. The predictive performance of the random forest was assessed using brier score on the validation data. The p-value for significant important variables was calculated based on Altmann et al. that depends on permutation importance [22].

A CART survival decision tree was constructed using the Rpart package; we used the minimum variable at each split of 10 and a maximum depth of 10 then we pruned the tree to avoid overfitting. Results were considered significant when the *p-value* was less than 0.05.

### Brier score

In this paper, the Brier score was used to compare the accuracy of prediction of each model. The Brier score measures the accuracy of the prediction. Brier score is an evaluation metric that calculates the weighted average of squared error between the event status at time t and predicted survival probability at time t. The higher the value of the Brier score, the less accurate the results are [23, 24]. The scores were calculated using the ipred package [25]

## Results

### Patients’ characteristics

The cohort included 26473 glioblastoma patients including training (n = 18538) and validation set (n = 7935). The median age of patients was 64 years old (Table 1). White males had the highest rates of glioblastoma. The most common site for glioblastoma was the frontal lobe. The median survival time for all patients was eight months. The cohort included patients who had anaplastic undifferentiated tumors (n = 7370). 77.4% of patients received surgical treatment; 93.4% of patients did not survive.

**Table 1.**
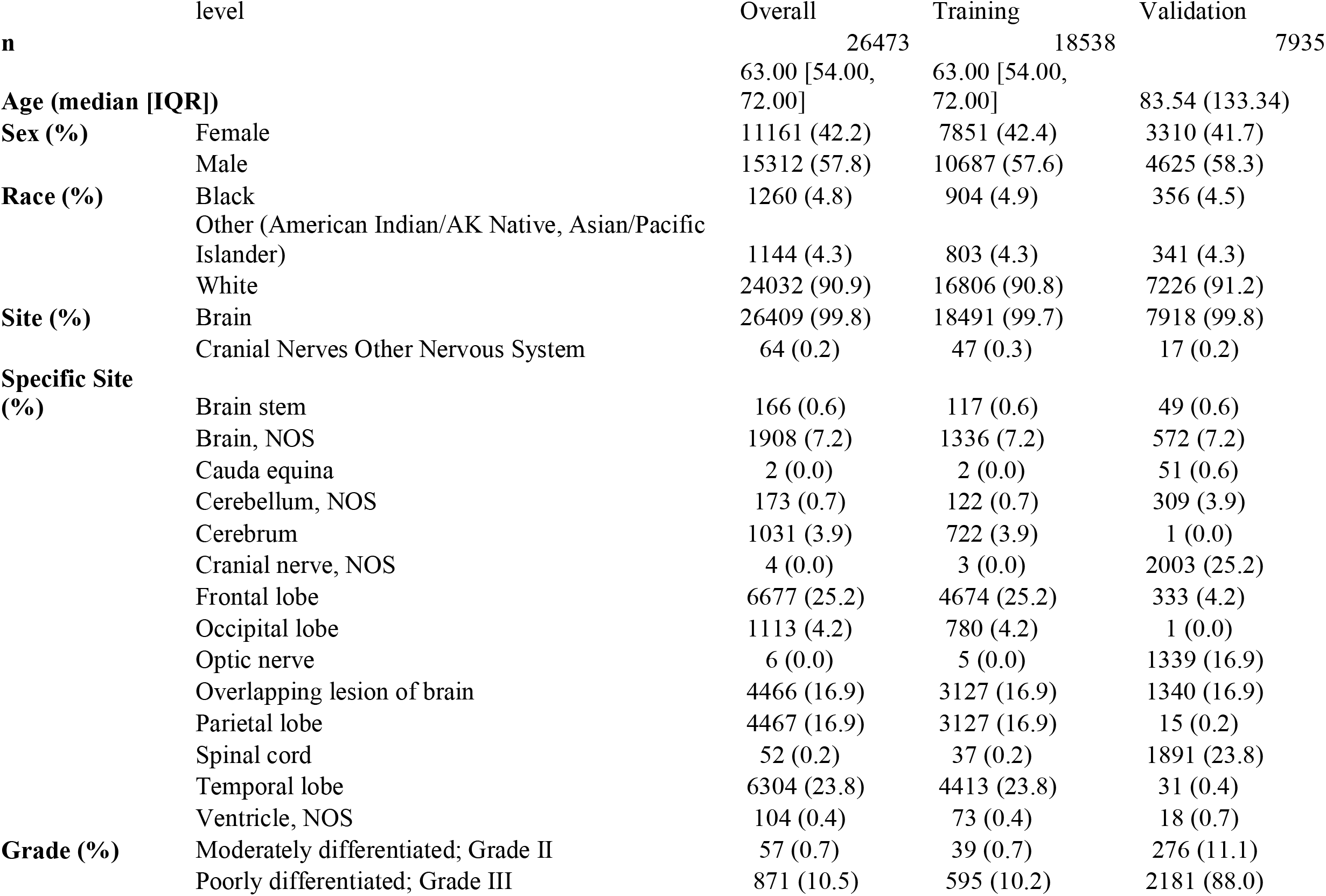
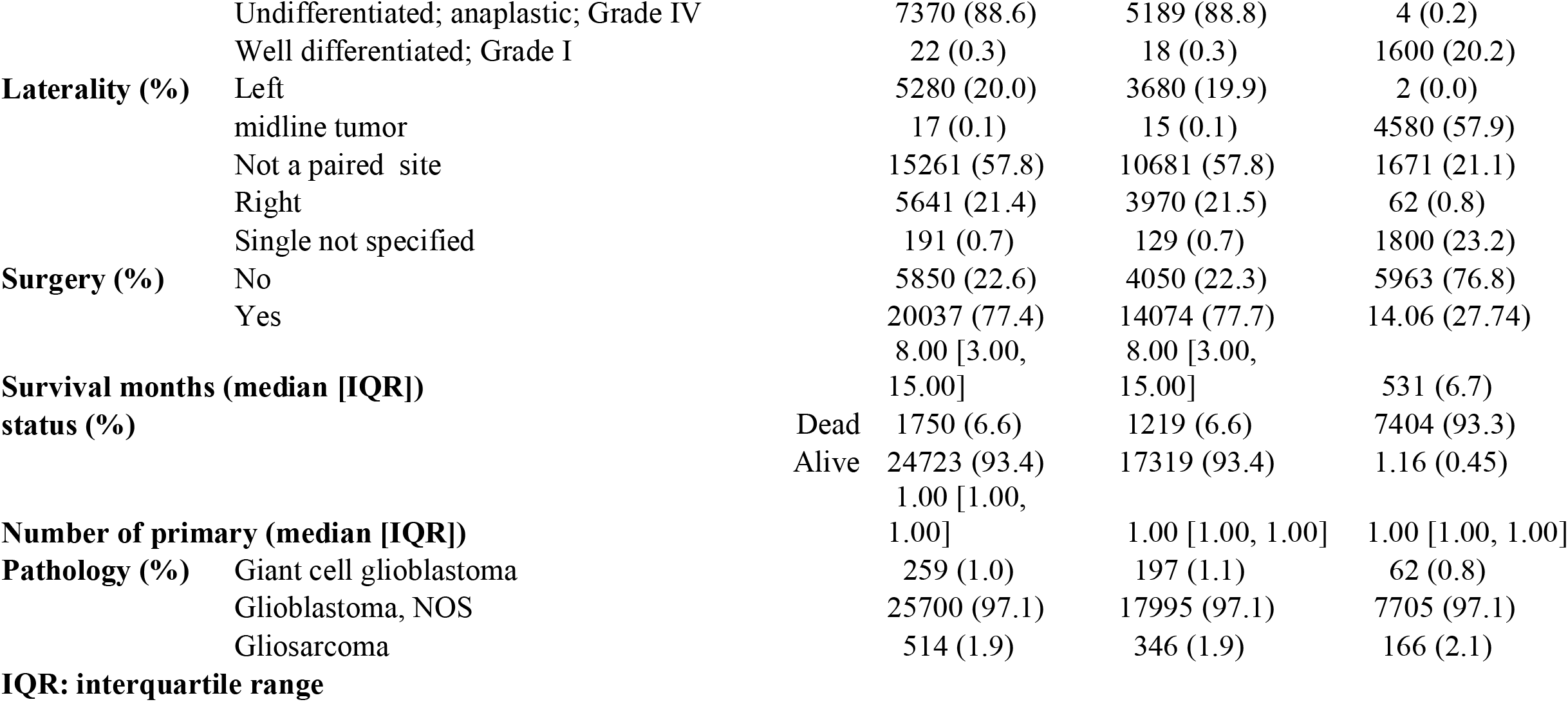
Characteristics of the included patients.

### Survival analysis using Cox Proportional Hazard regression (CPH)

The univariable analysis revealed that all patients characteristics except gender of the patients had increased or decreased the mortality rate, for instance, older age patients had low survival probability [HR = 1.2, 95% CI (0.87, 1.5), p-value <0.001]. For tumor characteristics, pathology, different sites of the tumor, Grade, and laterality had significantly affected the survival of glioblastoma patients (Table 2). In the multivariable analysis, old white patients were diagnosed with undifferentiated tumors in the optic nerve with more than one primary tumor (Table 2). Moreover, Gliosarcoma had decreased survival probability (Table 2).

**Table 2.** Univariable and multivariable cox proportional hazard model.

| Characteristic | N | HR<sup>/</sup> | 95%<br>CI<sup>/</sup> | p-value | HR<sup>/</sup> | 95%<br>CI<sup>/</sup> | p-value |
| --- | --- | --- | --- | --- | --- | --- | --- |
| **Age** | 18,538 | 1.2 | 0.87,<br>1.5 | <0.001 | 1 | 1.00,<br>1.00 | <0.001 |
| **Sex** | 18,538 |  |  |  |  |  |  |
| **Female** |  | — | — |  |  |  |  |
| **Male** |  | 0.98 | 0.95,<br>1.01 | 0.17 |  |  |  |
| **Race** | 18,538 |  |  |  |  |  |  |
| **Black** |  | — | — |  | — | — |  |
| **Other (American Indian/AK Native, Asian/Pacific Islander)** |  | 1.04 | 0.94,<br>1.15 | 0.47 | 1.05 | 0.95,<br>1.16 | 0.4 |
| **White** |  | 1.24 | 1.15,<br>1.33 | <0.001 | 1.22 | 1.13,<br>1.30 | <0.001 |
| **Site** | 18,538 |  |  |  |  |  |  |
| **Brain** |  | — | — |  | — | — |  |
| **Cranial Nerves Other Nervous System** |  | 0.71 | 0.52,<br>0.97 | 0.031 |  |  |  |
| **Specific Sites** | 18,538 |  |  |  |  |  |  |
| **Brain stem** |  | — | — |  | — | — |  |
| **Brain, NOS** |  | 1.58 | 1.30,<br>1.93 | <0.001 | 1.75 | 1.44,<br>2.14 | <0.001 |
| **Cauda equina** |  | 0.25 | 0.03,<br>1.79 | 0.17 | 0.35 | 0.05,<br>2.48 | 0.3 |
| **Cerebellum, NOS** |  | 0.98 | 0.75,<br>1.28 | 0.87 | 1.31 | 1.00,<br>1.71 | 0.049 |
| **Cerebrum** |  | 1.36 | 1.11,<br>1.67 | 0.003 | 1.68 | 1.37,<br>2.06 | <0.001 |
| **Cranial nerve, NOS** |  | 3.74 | 1.19,<br>11.8 | 0.024 | 3.33 | 1.06,<br>10.5 | 0.04 |
| **Frontal lobe** |  | 0.98 | 0.81,<br>1.19 | 0.84 | 1.53 | 1.26,<br>1.86 | <0.001 |
| **Occipital lobe** |  | 1.01 | 0.82,<br>1.24 | 0.92 | 1.56 | 1.27,<br>1.91 | <0.001 |
| **Optic nerve** |  | 2.07 | 0.84,<br>5.07 | 0.11 | 1.34 | 0.54,<br>3.29 | 0.5 |
| **Overlapping lesion of brain** |  | 1.29 | 1.06,<br>1.56 | 0.01 | 1.64 | 1.35,<br>1.99 | <0.001 |
| **Parietal lobe** |  | 1.07 | 0.88,<br>1.30 | 0.5 | 1.64 | 1.35,<br>1.99 | <0.001 |
| **Spinal cord** |  | 0.7 | 0.47,<br>1.04 | 0.08 | 0.88 | 0.59,<br>1.31 | 0.5 |
| **Temporal lobe** |  | 1.01 | 0.83,<br>1.22 | 0.94 | 1.59 | 1.31,<br>1.93 | <0.001 |
| **Ventricle, NOS** |  | 0.97 | 0.71,<br>1.31 | 0.83 | 1.3 | 0.96,<br>1.77 | 0.091 |
| **Grade** | 18,538 |  |  |  |  |  |  |
| **Moderately differentiated; Grade II** |  | — | — |  | — | — |  |
| **Poorly differentiated; Grade III** |  | 1.39 | 1.03,<br>1.88 | 0.03 | 1.4 | 1.04,<br>1.88 | 0.029 |
| **Undifferentiated; anaplastic; Grade IV** |  | 1.32 | 0.99,<br>1.77 | 0.061 | 1.44 | 1.08,<br>1.94 | 0.014 |
| **Well differentiated; Grade I** |  | 1.2 | 0.74,<br>1.93 | 0.46 | 1.42 | 0.88,<br>2.29 | 0.15 |
| **Laterality** | 18,538 |  |  |  |  |  |  |
| **Left** |  | — | — |  | — | — |  |
| **midline tumor** |  | 1.14 | 0.65,<br>2.01 | 0.65 | 1.17 | 0.66,<br>2.07 | 0.6 |
| **Not a paired site** |  | 1.48 | 1.42,<br>1.54 | <0.001 | 1.45 | 1.39,<br>1.52 | <0.001 |
| **Right** |  | 0.98 | 0.94,<br>1.03 | 0.53 | 1.02 | 0.97,<br>1.07 | 0.4 |
| **Single not specified** |  | 1.35 | 1.12,<br>1.62 | 0.001 | 1.2 | 0.99,<br>1.44 | 0.057 |
| **Surgery** | 18,538 |  |  |  |  |  |  |
| **No** |  | — | — |  | — | — |  |
| **Yes** |  | 0.47 | 0.45,<br>0.49 | <0.001 | 0.52 | 0.50,<br>0.54 | <0.001 |
| **numberofprimary** | 18,538 | 1.09 | 1.06,<br>1.13 | <0.001 | 1.08 | 1.05,<br>1.12 | <0.001 |
| **Pathology** | 18,538 |  |  |  |  |  |  |
| **Giant cell glioblastoma** |  | — | — |  | — | — |  |
| **Glioblastoma, NOS** |  | 1.42 | 1.22,<br>1.65 | <0.001 | 1.27 | 1.09,<br>1.48 | 0.002 |
| **Gliosarcoma** |  | 1.36 | 1.13,<br>1.64 | 0.001 | 1.44 | 1.20,<br>1.74 | <0.001 |
| <sup>†</sup>HR = Hazard Ratio, CI = Confidence Interval |  |  |  |  |  |  |  |

The accuracy of the model was checked using the Brier Score (BS) for both short- and long-term predictions. We found that the accuracy of the model was the least in the first year, followed by two years. After the sixth year, the model had the best accuracy that was sustained until 10-year predictions (Table 3). The same was found in the separate analysis with the validation set imputed through KNN. For machine learning algorithms.

**Table 3.** Comparison between each model using brier score to estimate the accuracy of each model for short- and long-term prediction in test set with or without missing data imputation

| **Time** | **Cox proportional Hazard ratio** |  | **CART** |  | **Random forest** |  |
| --- | --- | --- | --- | --- | --- | --- |
|  | **No imputation in the validation set** | **KNN imputation in the validation set** | **No imputation in the validation set** | **KNN Imputation in the validation set** | **No imputation in the validation set** | **KNN imputation in the validation set** |
| **12 months** | 0.563 | 0.658 | 1.541 | 1.609 | 0.177 | 0.173 |
| **24 months** | 0.321 | 0.392 | 1.740 | 1.801 | 0.105 | 0.152 |
| **36 months** | 0.245 | 0.309 | 1.787 | 1.851 | 0.058 | 0.126 |
| **48 months** | 0.220 | 0.279 | 1.802 | 1.866 | 0.0423 | 0.107 |
| **60 months** | 0.210 | 0.264 | 1.805 | 1.847 | 0.037 | 0.093 |
| **72 months** | 0.198 | 0.252 | 1.809 | 1.873 | 0.0303 | 0.08 |
| **84 months** | 0.192 | 0.243 | 1.812 | 1.883 | 0.072 | 0.075 |
| **96 months** | 0.188 | 0.239 | 1.814 | 1.886 | 0.066 | 0.069 |
| **108 months** | 0.184 | 0.230 | 1.815 | 1.889 | 0.060 | 0.064 |
| **120 months** | 0.181 | 0.226 | 1.817 | 1.889 | 0.019 | 0.059 |

Ensemble trees were constructed using 500 trees with three variables used at each split. The trees identified age, laterality, and surgery as the top three important variables for the prediction of patient survival (Figure 1). Random forest survival trees had high accuracy (low brier score); the BS was 0.177 for the first year and decreased for the long-term analysis (0.01 for 10 years’ prediction) Table 3.

**Figure 1.**
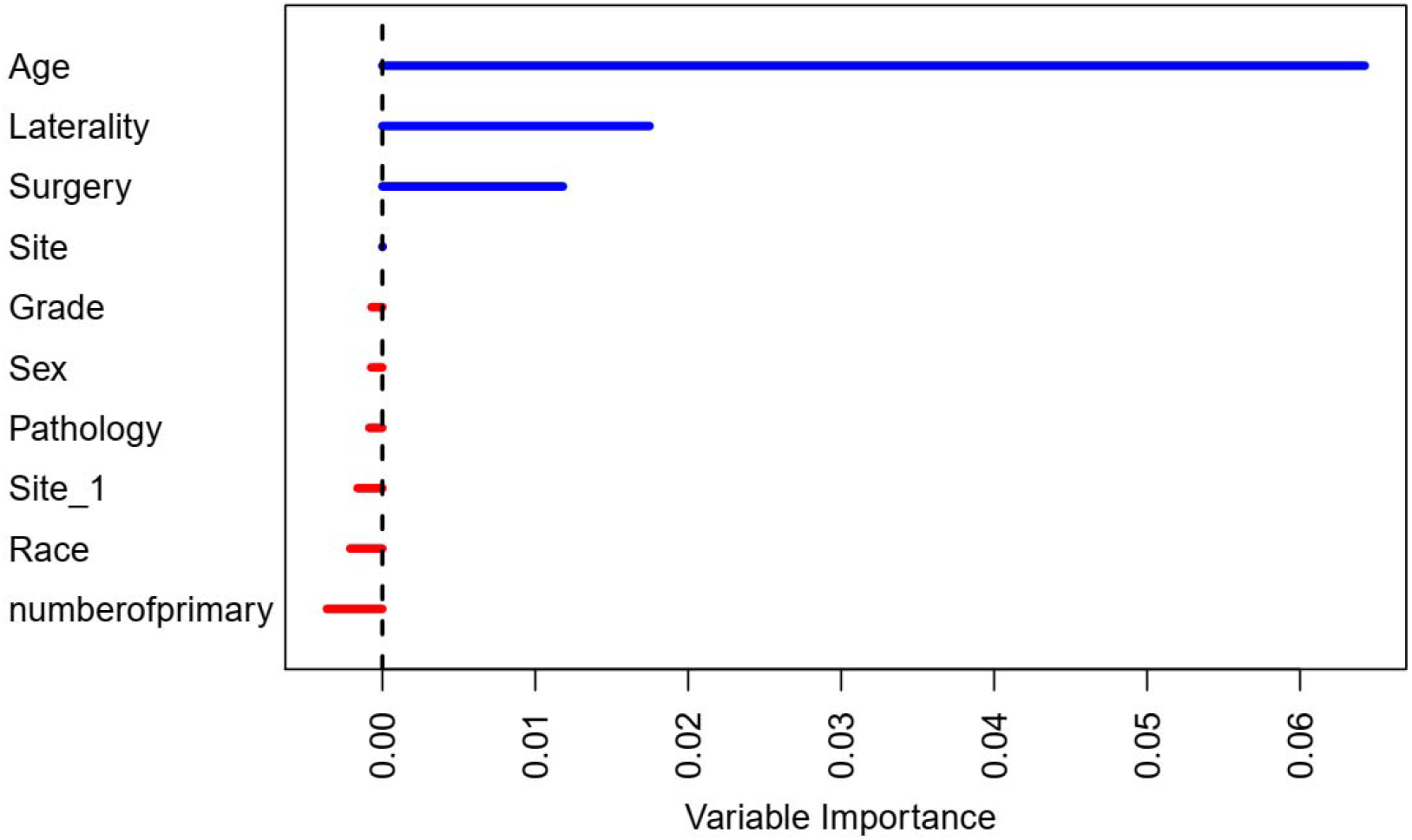
Variable importance plot illustrating the top variables used for prediction. Site_1 is the specific central nervous system sites.

For the CART survival tress that used all the predictors for survival prediction, the cohort was divided into six groups as shown in (Figure 2). The groups are: 1) Patients aged more than 66.5 and performed surgery (median survival = 6 months, n = 5079, 2) Patients aged more than 66.5 and did not perform any surgery (median survival = 3 months, n = 1872), 3) Patients who aged less than 45.5 (median survival = 16 months, n = 2283) have survival probability of 0.538, 4) Patients whose age are between 45.5 – 66.5 and performed surgery (median survival = 11 months, n = 7271), 5) Patients whose age are between 45.5 – 66.5 and did not perform surgery (median survival = 5 months, n = 1578)

**Figure 2.**
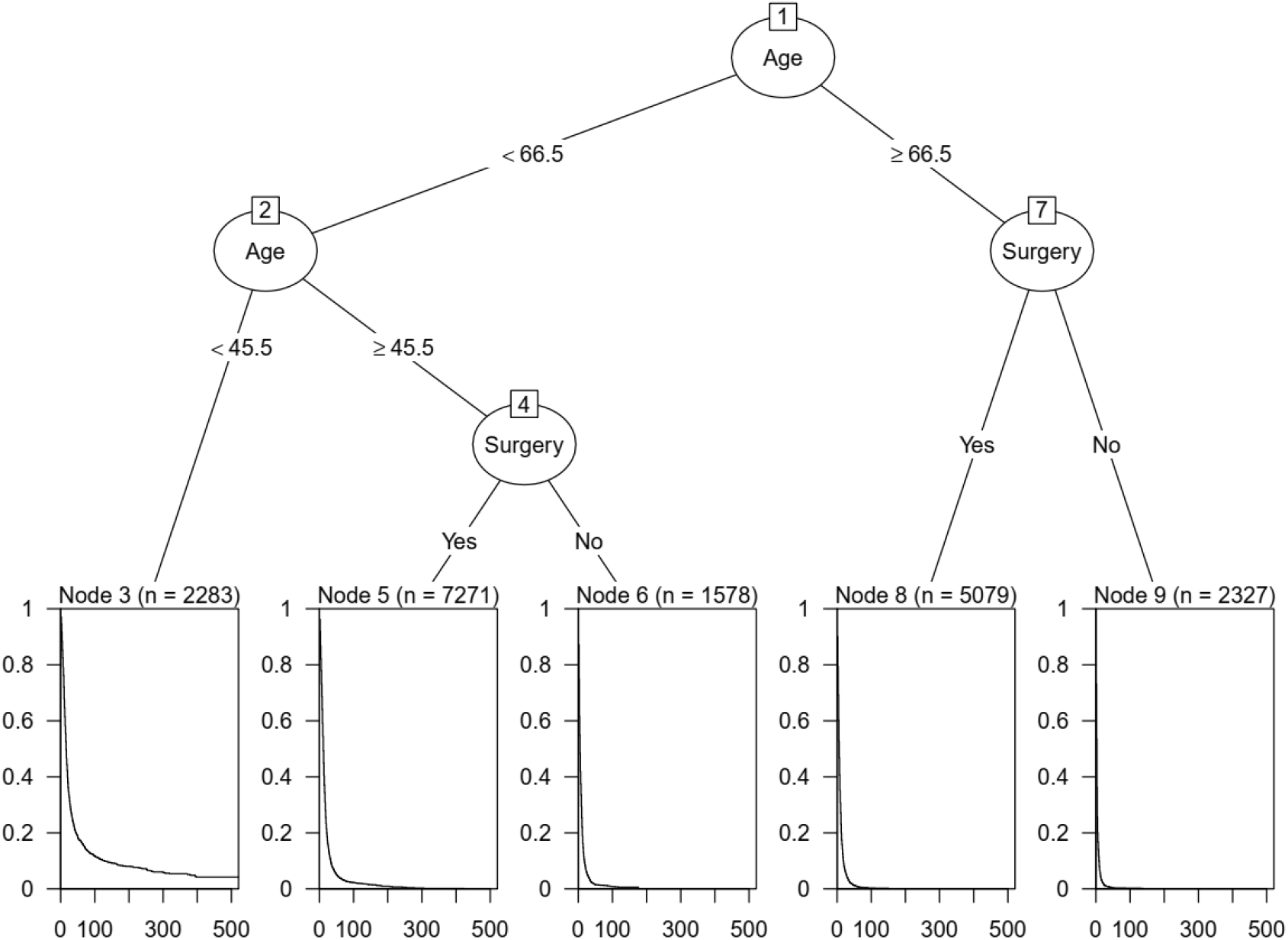
The recursive partitioning survival trees illustrating the survival probability for each group of patients.

Assessing the accuracy of the CART model using BS revealed that CART has much less accuracy than CPH and random forest implying that it is not suitable for long-term prediction Table 3. The accuracy of the model is decreasing through time with the highest accuracy in the first year (Figure 3).

**Figure 3.**
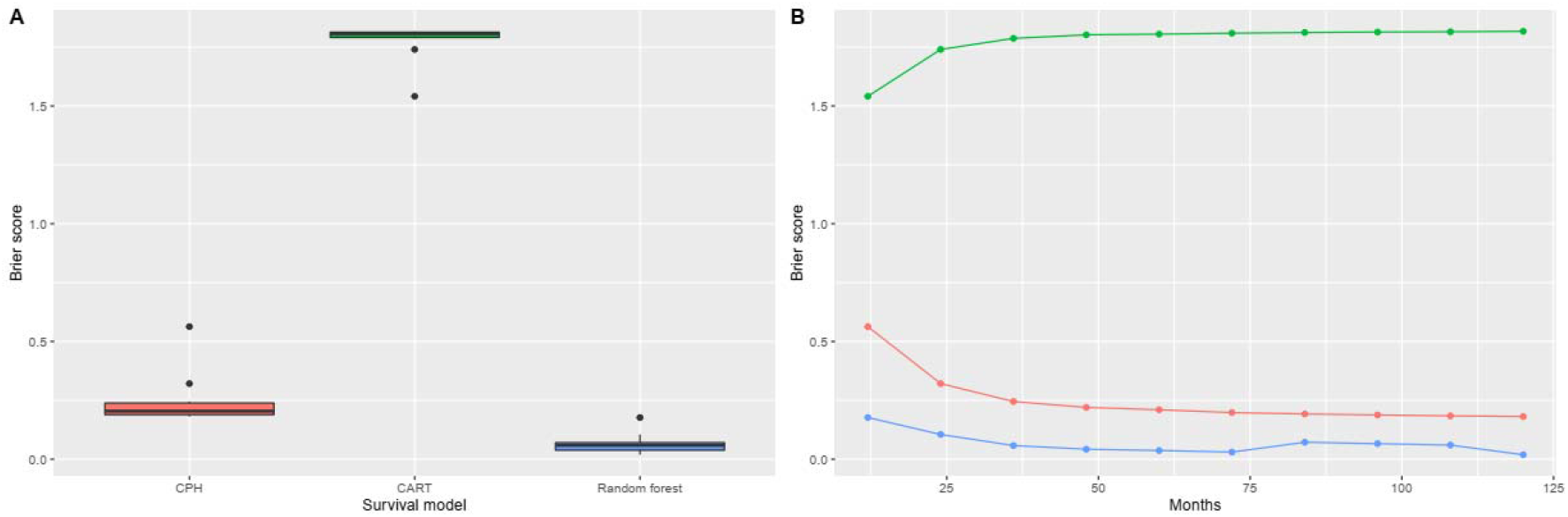
Boxplot showing the overall median brier score for each model suggesting random forest had the highest accuracy (A), Figure B shows the brier score at each time point indicating that random forest had the lowest brier score hence the highest accuracy

Based on our results, we found that random forest maintained high accuracy (low Brier score) for both short- and long-term predictions followed by CPH followed by CART survival trees Figure 3. Brier score for CPH model decreased for longer duration indicating the high accuracy of model for long term prediction, however, Brier score was higher than random forest suggesting random forest is the most suitable model for short- and long-term predictions Table 3. Recursive partitioning trees showed very low accuracy compared to other models.

## Discussion

This study aimed at investigating the appropriate survival models to use for the big data in medicine. We compared three models using Brier Scores (BS) as an indicator of model accuracy. Based on our results, RSF had the best accuracy for short- and long-term predictions followed by CPH.

Machine learning introduction into medical sciences and data analytics has created a massive impact in the field of public health [26]. With the help of automated processes and artificial intelligence, we acquired the ability to read unrevealed data and algorithm patterns.

According to Nasejje et al., when faced with irrelevance between the covariates in the model for time-to-event data, they use the Cox model and link each covariate with one public health assumption [27]. On the other hand, when using RSF, the data will be not reliant on the assumption for its validity [28]. So, Nasejje et al. used the RSF to analyze public health data to figure out associated factors with mortality of younger group of patients (under the age of five) [27]. Typically, in any dataset that includes many covariables carrying the risk of violating the proportional hazards assumption, RSF can be considered [29, 30].

Moreover, Random Survival Forests (RSF), in several risk models, outperformed the traditional Cox Proportional Hazard model (CPH). More importantly, while Cox cannot automatically identify the nonlinear effects of all considered variables, RSF can [28, 30-33]. However, in 2009, on breast cancer patients, the authors reported that CPH was a reliable method for predicting disease-free survival (DFS) in cancer. It was more advantageous than RSF approaches. This was justified by the capability of CPH to extract patterns and relationships hidden deep into medical datasets, leading to high predictive abilities that can be used for different sample sizes and potential future suitable survival data problems, whereas RSF provides only interpretive results [34].

For a dataset with separated and different risk levels, “Model-Based Recursive Partitioning” was able to present a good description. Safe M et al. reported the superiority of recursive partitioning for nonlinear model structures [35]. In the interaction dataset, recursive partitioning like Classification and Regression Trees (CART) and Artificial Neural Network (ANN) showed superior results to Cox (*P*< 0.05) with an improvement of 0.1 (95% CI, 0.08 to 0.12) and 0.015 (95% CI, 0.01 to 0.02) respectively. In theory, CART and ANN overcame the limitations of the Cox model regarding the ease and extent of their use [36]. In a study about breast tumor chemosensitivity to primary chemotherapy, on the other hand, the logistic regression model predictions were better than recursive partitioning [37, 38]. Another study by Lee et al. confirmed that Cox linear regression modeling outperformed recursive partitioning when there were only continuous predictors, while recursive partitioning was better when there were significant categorical predictors [37, 39]. One last study by Chun et al. demonstrated that Artificial Neural Network (ANN) had worse performance than the logistic regression model [37, 40]. Because the main goal of those methods (RSF, CART, and ANN) is to develop a predictive model of many variables that can lead to a more efficient clinical use that is particularly important for physicians. Contrastingly, conventional statistical modeling needs proper input of data from an expert to create a much easier model to interpret than data-driven-based techniques. This, in turn, leads to the narrow scope of conventional techniques to find new correlations between the data ready to be used in the literature [41].

Another study addressing dyslipidemia analyzed the difference in the disease incidence using RSF and Cox model and summarized that the RSF could predict more variables than the CPH. Those variables included the baseline lipoprotein profiles (including high- and low-density lipoproteins, total cholesterol, total triglycerides, blood pressure, age, body mass index, … etc.) [42]. Accordingly, not only do we need a tool to analyze the mortality, morbidity, and risk rates, but also a tool we can depend on searching for more variables effectively. So, using machine learning techniques can help us achieve combinations that we can barely capture using conventional approaches [41].

In our models, we validated our results using Brier Score (BS). An earlier study published in 2009 compared the RSF to CPH, using the Harrell c-statistics [43-45] for assessing the validity, found that CPH had better performance, and concluded that its replacement by RSF is still controversial and needed further investigation [46]. On the other hand, two other studies, assessing the 1-year mortality and survival in cardiac patients with cardiac arrhythmias, compared the use of RSF and the traditional CPH. The results were that RSF significantly overperformed CPH [47, 48]. The latter findings were supported by numerous other published studies [31, 42, 49, 50].

One of the biggest obstacles is not only to collect datasets that have the appropriate size and needed quality of samples but also to use appropriate methods in analysis. Towards that point, our study contributes as it compares these predictive methods which in turn can improve and reinforce the theory about the limitations and advantages of each method [26].

## Limitation

Our analyses were conducted in R. The effect of each level of the categorical variables in the survival trees could not be aptly illustrated due to the lack of R packages with more advanced illustrations. Another limitation is that random forest and the function used to calculate brier score failed if the data has missing values.

## Conclusion

In this paper, we compared the performance of the Cox Proportional Hazard model (CPH), Classification and Regression Trees (CART), and the Random Survival Forest (RSF) in predicting the survival of glioblastoma patients reported in the SEER database. We concluded that the RSF achieved the best performance and the highest accuracy followed by the CPH and lastly by the CART for both short- and long-term predictions, validated by the Brier Score (BS). Accordingly, using RSF may be of benefit in determining the best prognostic factors of cancer patients; however, more development of R packages is needed to allow for more illustrations of each covariate effect. More studies of the same kind are also needed to examine the performance of the three models in other cancer types.

## Data Availability

Availability of data and material
The data are available and can be accessed through the SEER database, which is publicly available at https://seer.cancer.gov/data/.

https://seer.cancer.gov/

## Declarations

### Ethics approval and consent to participate

Not applicable

### Consent for publication

The authors of this manuscript consent to the publication of the work by BioMed Central.

### Availability of data and material

The data are available and can be accessed through the SEER database, which is publicly available at https://seer.cancer.gov/data/.

### Authors’ Contributions

SM is responsible for the idea of the study, concept and design, acquisition of the data from the SEER database, statistical analysis and interpretation of data. SM, AMM, HT, OGH, NTMD, LDNN, and AHZ contributed to writing the manuscript. The study was under the supervision of NTH, who has also revised the manuscript. All authors read and approved the final version of the manuscript.

## Acknowledgments

None

## Code availability

The codes used in the analysis are available upon request from Dr. Sara Morsy

## Table legends

**Table 1**. The characteristics of included patients.

**Table 2**. Univariable and multivariable Cox regression analysis

**Table 3**. Comparison between each model using brier score for each model to estimate the accuracy of each model for short- and long-term prediction

